# Clinical Features of Patients Infected with the 2019 Novel Coronavirus (COVID-19) in Shanghai, China

**DOI:** 10.1101/2020.03.04.20030395

**Authors:** Min Cao, Dandan Zhang, Youhua Wang, Yunfei Lu, Xiangdong Zhu, Ying Li, Honghao Xue, Yunxiao Lin, Min Zhang, Yiguo Sun, Zongguo Yang, Jia Shi, Yi Wang, Chang Zhou, Yidan Dong, Longping Peng, Ping Liu, Steven M. Dudek, Zhen Xiao, Hongzhou Lu

## Abstract

**Background:** Since mid-December 2019, a cluster of pneumonia-like diseases caused by a novel coronavirus, now designated COVID-19 by the WHO, emerged in Wuhan city and rapidly spread throughout China. Here we identify the clinical characteristics of COVID-19 in a cohort of patients in Shanghai.

**Methods:** Cases were confirmed by real-time RT-PCR and were analysed for demographic, clinical, laboratory and radiological features.

**Results:** Of 198 patients, the median duration from disease onset to hospital admission was 4 days. The mean age of the patients was 50.1 years, and 51.0% patients were male. The most common symptom was fever. Less than half of the patients presented with respiratory systems including cough, sputum production, itchy or sore throat, shortness of breath, and chest congestion. 5.6% patients had diarrhoea. On admission, T lymphocytes were decreased in 45.8% patients. Ground glass opacity was the most common radiological finding on chest computed tomography. 9.6% were admitted to the ICU because of the development of organ dysfunction. Compared with patients not treated in ICU, patients treated in the ICU were older, had longer waiting time to admission, fever over 38.5° C, dyspnoea, reduced T lymphocytes, elevated neutrophils and organ failure.

**Conclusions:** In this single centre cohort of COVID-19 patients, the most common symptom was fever, and the most common laboratory abnormality was decreased blood T cell counts. Older age, male, fever over 38.5°C, symptoms of dyspnoea, and underlying comorbidity, were the risk factors most associated with severity of disease.

## Introduction

In December 2019, a cluster of patients with pneumonia of unknown cause was linked to a seafood wholesale market in Wuhan, China ^1^. Sequencing analysis from lower respiratory tract samples indicated a novel coronavirus, which was initially named 2019 novel coronavirus (2019-nCov), and later reclassified as SARS-CoV-2 by the World Health Organization. Coronaviruses are known to cause multiple system infections in various animals and primarily respiratory tract infections in humans, including recent outbreaks of severe acute respiratory syndrome (SARS) and Middle East respiratory syndrome (MERS) ^2^. Employing lessons learned from the SARS outbreak, extraordinary public health measures were launched to control spread of the SARS-CoV-2 virus. Beginning on Jan 23, 2020, local governments in Hubei Province including Wuhan, Huanggang etc., announced the suspension of public transportation, with closure of airports, railway stations, and city highways, to prevent further disease transmission. Despite these extensive efforts by the Chinese government and health officials to control the outbreak of coronavirus disease 2019 (COVID*-*19), by January 30th, 2020, human-to-human transmission had been reported to occur outside Wuhan, extending into other regions of China and to other countries. WHO has declared that the outbreak of COVID-19 constitutes a Public Health Emergency of International Concern (PHEIC). As of Feb 19, 2020, 72,533 confirmed cases had been reported with a total of 1872 deaths.

At the time of this analysis, only a little over a month has passed since COVID-19 was first reported ^1^. There are reasonable concerns about whether COVID-19 has undergone rapid or marked genomic mutation during transmission. Yet large cohort reports from outside Wuhan (Hubei Province) are not yet available, and the clinical characteristics of COVID-19 remain largely unclear. In this article, we describe the initial clinical, laboratory, and radiological characteristics of patients confirmed to have COVID-19 in Shanghai, and we compare the clinical features between patients with less severe illness and those requiring critical care.

## Methods

### Patients

We obtained epidemiological, demographic, clinical, laboratory and management data from the medical records of patients infected with SARS-Cov-2. On Jan 20, 2020, the first human case of COVID-19 in Shanghai was confirmed. Since then all hospitals in Shanghai have opened special fever clinics to screen suspected patients, and laboratory confirmed patients were then admitted to a single designated hospital in Shanghai (Shanghai Public Health Clinical Centre). Laboratory confirmation of COVID-19 was done by the Chinese Centre for Disease Control and Prevention. Throat-swab specimens from the upper respiratory tract were obtained from all patients at admission and maintained in viral transport medium. COVID-19 was confirmed by real-time RT-PCR using the same protocol as described previously ^3^. Confirmed patients were hospitalized into negative pressure wards for further medical observation and treatment. We collected data from patients who were admitted from Jan. 20 up to Feb. 15. All the data collected from the included cases have been shared with the WHO.

### Data Collection

Epidemiological exposure data, patient characteristics, clinical symptoms, laboratory and imaging findings and medical history were extracted from electronic medical records and analysed by licensed physicians. Laboratory data were recorded in standardized form. Initial investigations included a complete blood count, routine urinalysis, blood gases, coagulation function, erythrocyte sedimentation rate (ESR), C-reactive protein (CRP) and serum biochemical testing (including renal and liver function, serum lactate, lactate dehydrogenase, and electrolytes). To characterize the effect of coronavirus on the immune system, immunologic factors including serum immunoglobulin, complements, cytokines, rheumatoid factor (RF) and T cell and leukocyte subpopulations were analysed by flow cytometry.

Non-laboratory information, including patient characteristics, epidemiological and medical history, were obtained and recorded by licensed physicians from direct interviews of patients and their relatives. Two researchers independently reviewed the data collection forms to review the data extracted. Written informed consent was waived in light of the urgent need to collect clinical data.

The severity of COVID was defined based on the criteria established by China’s National Health Commission ^4^. 1. Mild. Minor symptoms only, without evidence for pneumonia by chest X-ray. 2, Moderate. Fever and respiratory symptoms are present, and there is evidence for pneumonia by chest X-ray. 3. Severe. Defined by any of the following conditions. 1) Dyspnoea, respiratory rate ≥30 /min, 2) resting hypoxia SaO2≤93%, 3) PaO2/FiO2 ≤300 mmHg. 4. Critical. The presence of any of the following conditions. 1) Respiratory failure, require mechanical ventilation, 2) shock, 3) other acute organ failure.

### Statistical analysis

Median and interquartile range (IQR) or mean and standard deviation were calculated for continuous variables. Count and percentages were presented for categorical variables. Statistical Analysis was performed by using SPSS21.0.

## Results

### Demographic and clinical characteristics

The demographic and clinical characteristics are shown in Table 1. Of 198 confirmed cases, the mean age of the patients was 50.1 years (± 16.3). 48.5% of the patients were under 50 years old, among them 66 (33.3%) were under 39 years old, and 30 were aged 40-49. 24 (12.1%) patients were over 70 years old. 14 (7.1%) of the patients had a history of alcohol use, and 11 (5.6%) had a smoking history. 101 (51.0%) patients were male, and 97 (49.0%) were female. 69 patients had at least one underlying chronic medical disorder. These included 42 (21.2%) patients with hypertension, 15 (7.6%) with diabetes, 12 (6.0%) with cardiovascular diseases, 4 (2.0%) with malignancy, and 6 (3.0%) with thyroid diseases.

**Table 1:** Demographics and baseline characteristics of patients with COVID-19

|  | Patients (n=198) | ICU(n=19) | Non-ICU(n=179) | P value |
| --- | --- | --- | --- | --- |
| Time between symptom onset and hospital admission, days | 4.0 (2.0-7.0) | 7.0 (5.0-9.0) | 4.0 (2.0-7.0) | 0.03 |
| Characteristics |  |  |  |  |
| Age, years | 50.1 (16.3) | 63.7 (16.8) | 48.6 (15.6) | <0.001 |
| $\leq 39$ | 66 (33.3%) | 2 (10.5%) | 64 (35.8%) | - |
| 40-49 | 30 (15.2%) | 2 (10.5%) | 28 (15.6%) | - |
| 50-59 | 33 (16.7%) | 0 (0.0%) | 33 (18.4%) | - |
| 60-69 | 45 (22.7%) | 7 (36.8%) | 38 (21.2%) | - |
| $\geq 70$ | 24 (12.1%) | 8 (42.1%) | 16 (8.9%) | - |
| Sex |  |  |  |  |
| Male | 101 (51.0%) | 17 (89.5%) | 84 (46.9%) | <0.001 |
| Female | 97 (49.0%) | 2 (10.5%) | 95 (53.1%) |  |
| Exposure History to Hubei Province | 126 (63.6%) | 9 (47.4%) | 117 (65.4%) | 0.121 |
| Smoking | 11 (5.6%) | 1 (5.3%) | 10 (5.6%) | 0.953 |
| Alcohol Use | 14 (7.1%) | 1 (5.3%) | 13 (7.3%) | 0.746 |
| **Any comorbidity** | **69 (34.8)** | **10 (52.6%)** | **59 (33.0%)** | **0.872** |
| Hypertension | 42 (21.2%) | 6 (31.6%) | 36 (20.1%) | 0.245 |
| Diabetes | 15 (7.6%) | 2 (10.5%) | 13 (7.3%) | 0.609 |
| Cardiovascular disease | 12 (6.0%) | 5 (26.3%) | 7 (3.9%) | <0.001 |
| Liver disease | 6 (3.0%) | 0 (0.0%) | 6 (3.4%) | 1.000 |
| Malignancy | 4 (2.0%) | 0 (0.0%) | 4 (2.2%) | 1.000 |
| Thyroid Disease | 6 (3.0%) | 1 (5.3%) | 5 (2.8%) | 0.459 |
| **Signs and symptoms** |  |  |  |  |
| Fever | 172 (86.9%) | 18 (94.7%) | 154 (86.0%) | 0.286 |
| Highest temperature, °C | 38.3 (38-38.9) | 38.95 (38.425-39.05) | 38.1 (37.8-38.6) | 0.002 |
| < 38.4 | 91 (58.7%) | 3 (21.4%) | 88 (62.4%) | <0.001 |
| > 38.5 | 64 (41.3%) | 11 (78.6%) | 53 (37.6%) | - |
| Cough | 97 (50.0%) | 7 (36.8%) | 90 (50.3%) | 0.265 |
| Sputum production | 55 (27.8%) | 5 (26.3%) | 50 (27.9%) | 0.881 |
| Itchy or sore throat | 19 (9.6%) | 0 (0.0%) | 19 (10.6%) | 0.135 |
| Myalgia or fatigue | 59 (29.8%) | 6 (31.6%) | 53 (29.6%) | 0.858 |
| Headache or dizziness | 24 (12.1%) | 0 (0.0%) | 24 (13.4%) | 0.089 |
| Diarrhoea | 11 (5.6%) | 0 (0.0%) | 11 (6.1%) | 0.266 |
| Chest congestion | 16 (8.1%) | 3 (15.8%) | 13 (7.3%) | 0.159 |
| Shortness of breath | 11 (5.6%) | 7 (36.8%) | 4 (2.2%) | <0.001 |

The most common symptom identified was fever (86.9%). Patients’ body temperatures were most commonly in a range from 38-38.9 °C (IQR). 10 (8.8%) patients presented with high fever greater than 39[. Less than half of the patients presented with respiratory systems including cough (46.4%), sputum production (23.2%), itchy or sore throat (9.8%), shortness of breath (4.5%) and chest congestion (8.0%). Myalgia or fatigue occurred in almost one-third (31.3%) of the patients. In addition, 5 (4.4%) patients had diarrhoea, which is suggestive of digestive system involvement (Table 1).

No infections of health care workers were detected during the time interval studied. A history of recent travel to Wuhan, and contact with people from Wuhan was documented in 126 (63.6%) of the patients. There was no difference in severity of disease between cases with Wuhan contact history and no-Wuhan contact history.

Of these 198 patients, 179 (90.4%) were admitted to isolation wards, including 3 mild and 176 moderate cases. 19 (9.5%) were admitted and transferred to the ICU because of the development of respiratory failure or other organ dysfunction, including 9 severe cases and 10 critical cases. 1 patient died without collecting sufficient data, and therefore was excluded from this study. Patient age differed significantly between the two groups (63.7 ± 16.8 for ICU vs 48.6 ± 15.6 for non-ICU patients). Male patients were significantly more common in ICU as compared with non-ICU cases (89.5% vs. 46.9%). Moreover, patients with underlying cardiovascular diseases were significantly more common in ICU cases as compared with non-ICU cases (26.3% vs. 3.9%, P<0.01). Compared with the non-ICU, patients admitted to the ICU were more likely to have high fever with temperature over 38.5°C (78.6% vs 37.6%), shortness of breath (36.8% vs 2.2%), and longer waiting period from onset of symptom to hospital admission (7 vs 4 days).

### Blood cell counts, coagulation function and other laboratory findings

On admission, the majority of patients had normal white blood cell, neutrophil, lymphocyte and platelet counts (Table 2). Prothrombin time (PT) (median 13.3, 12.9-13.8) and activated partial thromboplastin time (APTT, median 39.5, 36.7-42.8) were normal in most patients. However, a marked rise in fibrinogen and CRP was observed in 108 (55.4%) and 152 (78.4%) cases, respectively. The majority of patients (65.4%) had decreased blood calcium concentration, while 19.5% cases had decreased blood sodium concentration.

**Table 2:**
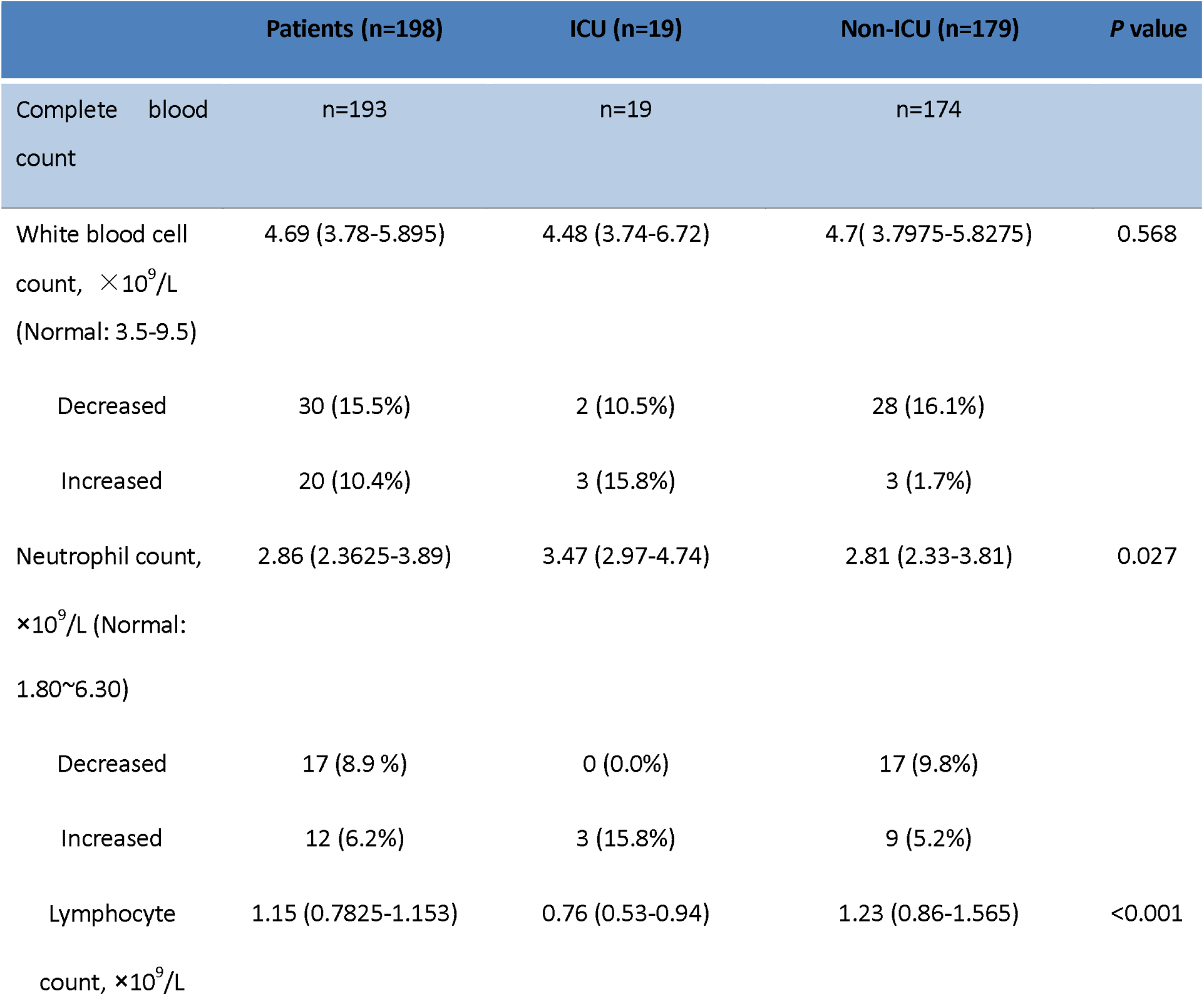
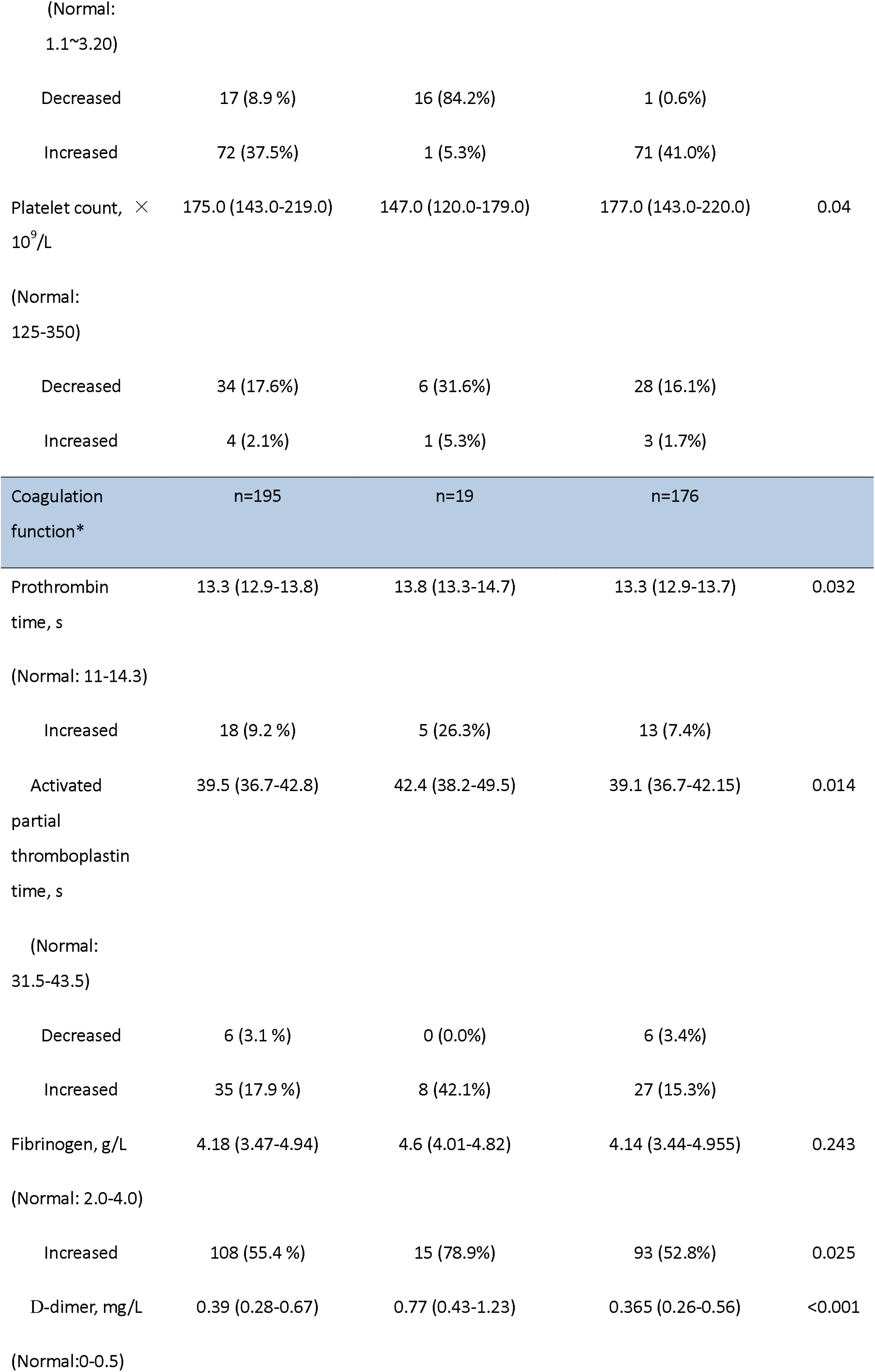
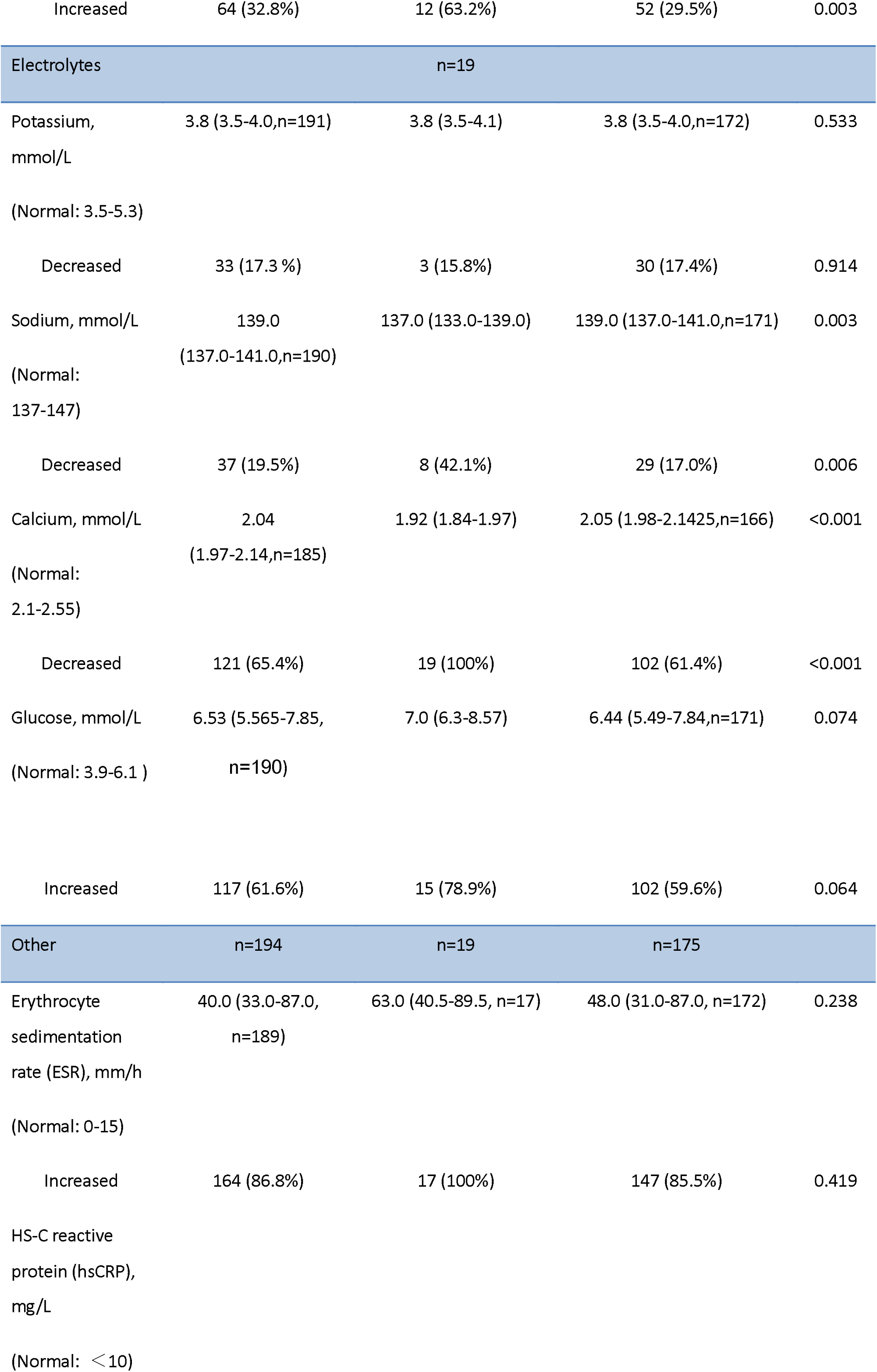
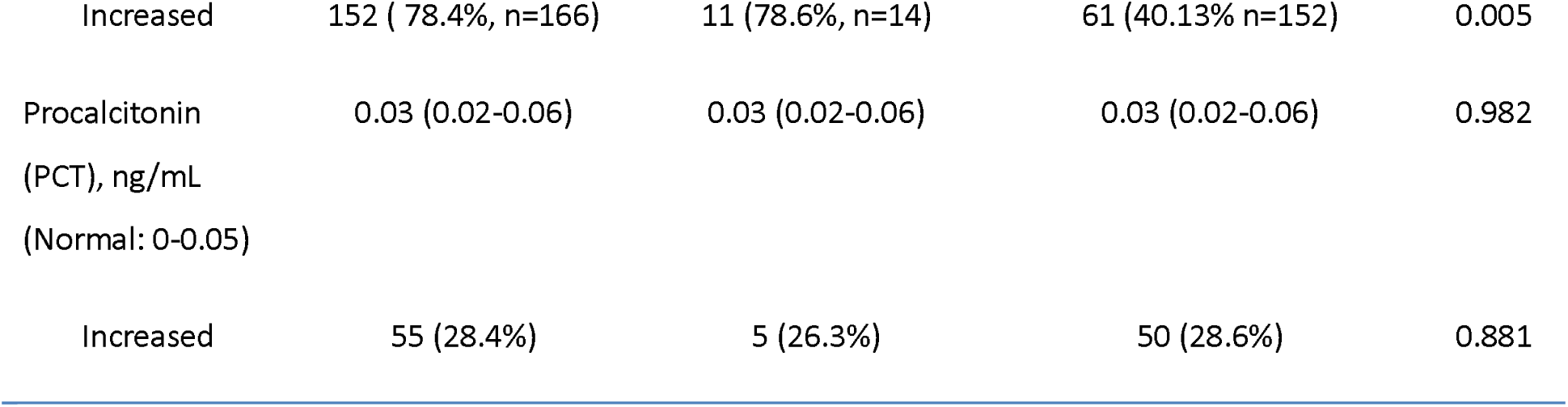
Admission blood cell count and coagulation function of patients with COVID-19

|  | Patients (n=198) | ICU (n=19) | Non-ICU (n=179) | P value |
| --- | --- | --- | --- | --- |
| Complete blood count | n=193 | n=19 | n=174 |  |
| White blood cell count, $\times 10^9/L$ (Normal: 3.5-9.5) | 4.69 (3.78-5.895) | 4.48 (3.74-6.72) | 4.7 (3.7975-5.8275) | 0.568 |
| Decreased | 30 (15.5%) | 2 (10.5%) | 28 (16.1%) |  |
| Increased | 20 (10.4%) | 3 (15.8%) | 3 (1.7%) |  |
| Neutrophil count, $\times 10^9/L$ (Normal: 1.80~6.30) | 2.86 (2.3625-3.89) | 3.47 (2.97-4.74) | 2.81 (2.33-3.81) | 0.027 |
| Decreased | 17 (8.9%) | 0 (0.0%) | 17 (9.8%) |  |
| Increased | 12 (6.2%) | 3 (15.8%) | 9 (5.2%) |  |
| Lymphocyte count, $\times 10^9/L$ | 1.15 (0.7825-1.153) | 0.76 (0.53-0.94) | 1.23 (0.86-1.565) | <0.001 |

|  |  |  |  |
| --- | --- | --- | --- |
| Decreased | 17 (8.9 %) | 16 (84.2%) | 1 (0.6%) |
| Increased | 72 (37.5%) | 1 (5.3%) | 71 (41.0%) |

|  |  |  |  |
| --- | --- | --- | --- |
| Decreased | 34 (17.6%) | 6 (31.6%) | 28 (16.1%) |
| Increased | 4 (2.1%) | 1 (5.3%) | 3 (1.7%) |

| Coagulation<br>function* | n=195 | n=19 | n=176 |
| --- | --- | --- | --- |

|  |  |  |  |
| --- | --- | --- | --- |
| Increased | 18 (9.2 %) | 5 (26.3%) | 13 (7.4%) |

|  |  |  |  |
| --- | --- | --- | --- |
| Decreased | 6 (3.1 %) | 0 (0.0%) | 6 (3.4%) |
| Increased | 35 (17.9 %) | 8 (42.1%) | 27 (15.3%) |
| Increased | 108 (55.4 %) | 15 (78.9%) | 93 (52.8%) | 0.025 |
| Increased | 64 (32.8%) | 12 (63.2%) | 52 (29.5%) | 0.003 |
| Electrolytes |  | n=19 |  |  |
| Potassium, mmol/L | 3.8 (3.5-4.0,n=191) | 3.8 (3.5-4.1) | 3.8 (3.5-4.0,n=172) | 0.533 |
| (Normal: 3.5-5.3) |  |  |  |  |
| Decreased | 33 (17.3 %) | 3 (15.8%) | 30 (17.4%) | 0.914 |
| Sodium, mmol/L | 139.0 | 137.0 (133.0-139.0) | 139.0 (137.0-141.0,n=171) | 0.003 |
| (Normal: 137-147) | (137.0-141.0,n=190) |  |  |  |
| Decreased | 37 (19.5%) | 8 (42.1%) | 29 (17.0%) | 0.006 |
| Calcium, mmol/L | 2.04 | 1.92 (1.84-1.97) | 2.05 (1.98-2.1425,n=166) | <0.001 |
| (Normal: 2.1-2.55) | (1.97-2.14,n=185) |  |  |  |
| Decreased | 121 (65.4%) | 19 (100%) | 102 (61.4%) | <0.001 |
| Glucose, mmol/L | 6.53 (5.565-7.85, n=190) | 7.0 (6.3-8.57) | 6.44 (5.49-7.84,n=171) | 0.074 |
| (Normal: 3.9-6.1 ) |  |  |  |  |
| Increased | 117 (61.6%) | 15 (78.9%) | 102 (59.6%) | 0.064 |
| Other | n=194 | n=19 | n=175 |  |
| Erythrocyte sedimentation rate (ESR), mm/h | 40.0 (33.0-87.0, n=189) | 63.0 (40.5-89.5, n=17) | 48.0 (31.0-87.0, n=172) | 0.238 |
| (Normal: 0-15) |  |  |  |  |
| Increased | 164 (86.8%) | 17 (100%) | 147 (85.5%) | 0.419 |
| HS-C reactive protein (hsCRP), mg/L |  |  |  |  |
| (Normal: <10) |  |  |  |  |
| Increased | 152 ( 78.4%, n=166) | 11 (78.6%, n=14) | 61 (40.13% n=152) | 0.005 |
| Procalcitonin<br>(PCT), ng/mL<br>(Normal: 0-0.05) | 0.03 (0.02-0.06) | 0.03 (0.02-0.06) | 0.03 (0.02-0.06) | 0.982 |
| Increased | 55 (28.4%) | 5 (26.3%) | 50 (28.6%) | 0.881 |

Compared with the non-ICU patients, patients admitted to the ICU were more likely to have increased neutrophil count (15.8% vs 5.2%), decreased lymphocyte count (84.2% vs 0.6%), increased prothrombin time (26.3% vs 7.4%), increased APTT (42.1% vs 15.3), increased fibrinogen (78.9% vs 52.8%) and increased D-dimer (63.2% vs 29.5%), as well as decreased levels of blood sodium (42.1% vs 17.0%) and calcium (100% vs 61.4%). By contrast, patients admitted to the ICU were less likely to have increased C-reactive protein (57.9% vs 80.6%).

### Organ dysfunction

The organ dysfunctions of the 198 patients are shown in Table 3. 21 (10.8%) patients had increased alanine aminotransferase (ALT) and 26 (20.0%) had increased aspartate aminotransferase (AST). Although 78 (40.0%) patients presented with hypoalbuminemia, the median (40.92, 37.99 - 43.12) was close to the normal range. Several patients had varying degrees of renal dysfunction or damage, with elevated blood urea nitrogen (15, 7.9%) or serum creatinine (10, 5.3%). 36.4% of the patients had positive urine protein tests. 22 (11.3%) patients had increased cardiac troponin I, and 33 (17.0%) patients had increased myoglobin.

**Table 3:** Organ dysfunction of patients with COVID-19 on admission to hospital

| Liver Function | n=195 | n=19 | n=176 |  |
| --- | --- | --- | --- | --- |
| Alanine<br>aminotransferase(ALT),<br>U/L<br>(Normal:9-50) | 23.0 (15.0-33.0) | 30.0 (19.0-34.0) | 22.0 (15.0-32.75) | 0.049 |
| Increased | 21 (10.8%) | 3 (15.8%) | 18 (10.2%) | 0.440 |
| Aspartate<br>aminotransferase(AST),<br>U/L<br>(Normal:9-40) | 26.0 (20.0-34.0) | 33.0 (26.0-51.0) | 24.0 (19.0-33.0) | <0.001 |
| Increased | 34 (17.4 %) | 8 (42.1%) | 26 (14.8%) | 0.002 |
| Total bilirubin(TB),<br>μmol/L<br>(Normal: 3.4-20.5) | 8.1 (6.5-10.6) | 9.0 (7.6-13.0) | 8.0 (6.5-10.35) | 0.013 |
| Increased | 5 (2.6%) | 1 (5.26%) | 4 (2.3%) | 0.399 |
| Albumin(ALB), g/L<br>(Normal:40-55) | 40.92<br>(37.99-43.12) | 37.17 (33.26-39.61) | 41.14 (38.83-43.3975) | <0.001 |
| Decreased | 78 (40.0 %) | 16 (84.2%) | 63 (35.8%) | <0.001 |
| Glutathione<br>reductase(GR), U/L<br>(Normal: 33-73) | 69.265<br>(63.3175-80.3625)<br>(n=194) | 86.12<br>(69.5525-107.6375)(n=18) | 68.75 (62.23-77.965) | <0.001 |
| Increased | 78 (40.2%) | 12 (66.7%) | 65 (36.9%) | 0.006 |
| Kidney function | n=190 | n=19 | n=171 |  |
| Creatinine(Cr), μmol/L<br>(Normal: 57-97) | 63.24<br>(51.695-76.9925) | 73.63 (56.96-84.55) | 62.7 (50.48-75.46) | 0.009 |
| Increased | 10 (5.3%) | 3 (15.8%) | 7 (4.1%) | 0.025 |
| Blood Urea<br>Nitrogen(BUN),<br>mmol/L<br>(Normal: 3.1-8.0) | 4.485<br>(3.5375-5.6025) | 4.54 (3.53-8.23) | 4.47 (3.54-5.57) | 0.463 |
| Increased | 15 (7.9%) | 5 (26.3%) | 10 (5.8%) | 0.001 |
| Serum lactate(LACT),<br>mmol/L<br>(Normal: 0.5-2.2) | 2.76 (2.36-3.175) | 2.57 (2.25-3.05) | 2.77 (2.36-3.21) | 0.506 |
| Increased | 159 (83.7%) | 16 (84.2%) | 143 (83.6%) | 0.652 |
| Urine Protein ($\geq 1+$) | 59 (36.4%,n=162) | 8 (44.4%,n=18) | 51 (35.4% ,n=144) | 0.217 |
| Myocardial enzymes | n=194 | n=19 | n=175 |  |
| Cardiac troponin I (cTnI), ng/mL | 0.02 (0.011-0.0385) | 0.039 (0.02-0.09) | 0.019 (0.011-0.0353) | 0.001 |
| (Normal: 0-0.04) |  |  |  |  |
| Increased | 22 (11.3 %) | 9 (47.4%) | 13 (7.4%) | <0.001 |
| Myoglobin, ng/mL | 5.92 (2.76-15.73) | 32.52 (17.4-70.17) | 4.89 (2.49-13.32) | <0.001 |
| (Normal: 0-48.8) |  |  |  |  |
| Increased | 33 (17.0 %) | 6 (31.6%) | 27 (15.3%) | 0.075 |

Compared with the non-ICU patients, patients admitted to the ICU were more likely to have increased AST (42.1% vs 14.8%), decreased albumin (84.2% vs 35.8%), and increased glutathione reductase (66.7% vs 36.9%). Patients admitted to the ICU were also more likely to have increased creatinine (15.8% vs 4.1%) and blood urea nitrogen (26.3% vs 5.7%) as measures of kidney dysfunction, as well as increased level of blood cardiac troponin I (47.4% vs 7.4%) indicative of heart injury.

### Immunological dysfunction

Most patients had decreased T lymphocyte counts, with 88 (45.8%) having decreased CD3+ T lymphocytes, 88 (45.8%) having decreased CD4 Th1 cells, 66 (34.4%) having decreased CD8 Th2 cells, and 73 (38.0%) having decreased CD45 positive cells, a pan-leukocyte marker. The majority of patients showed normal CD4/CD8 ratio, and normal levels of IgA, IgG, IgM, C3 and C4 (Table 4).

**Table 4:**
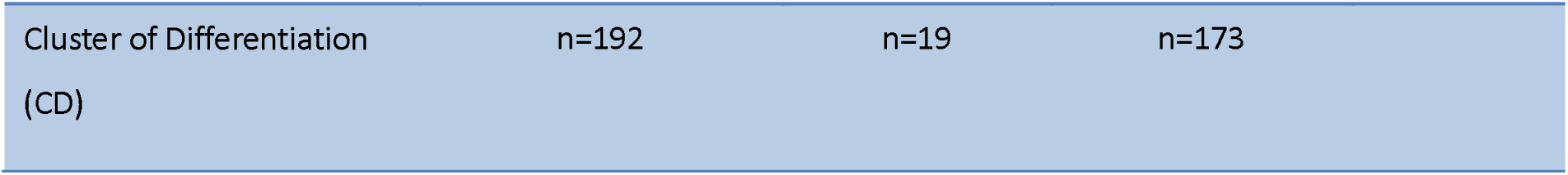
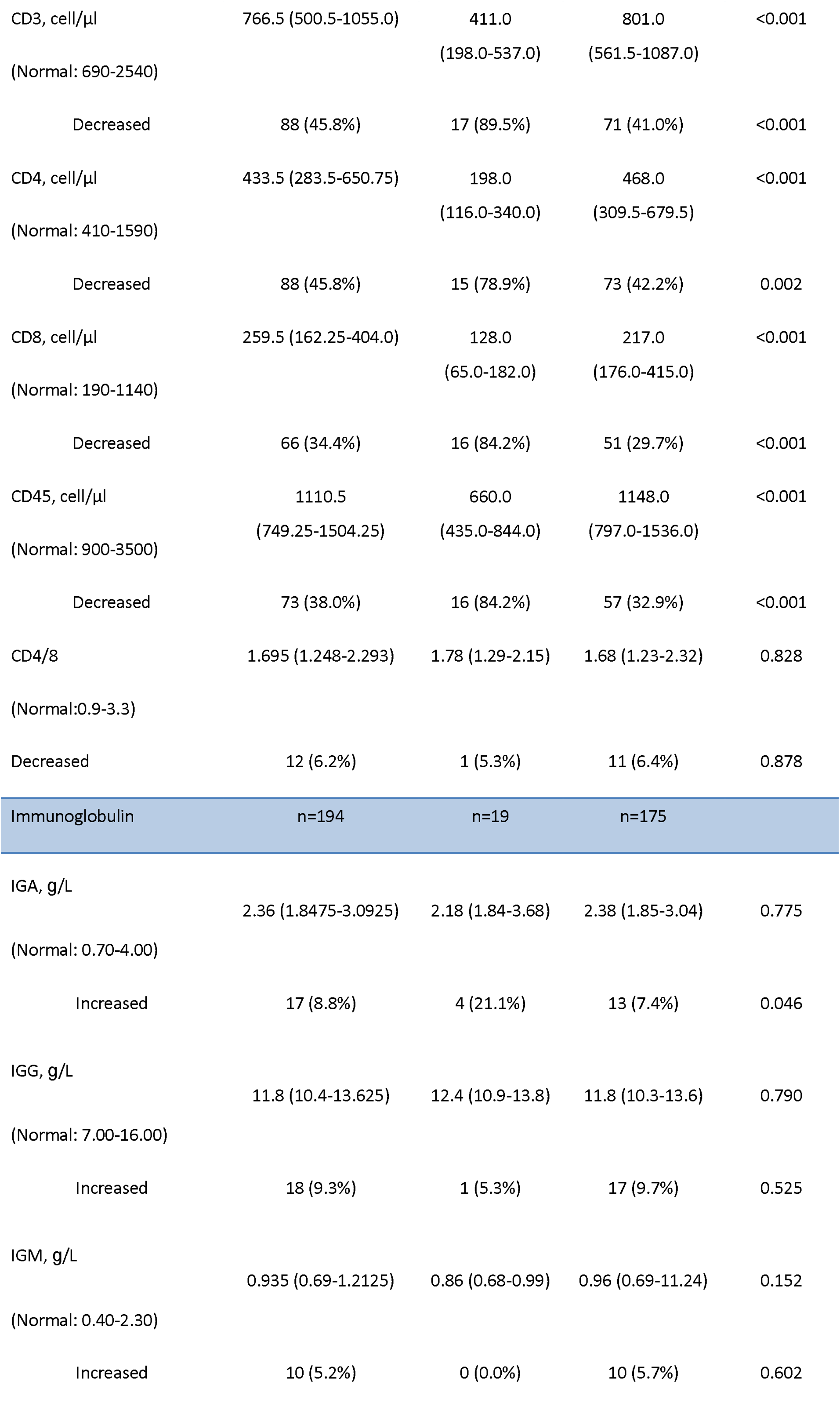
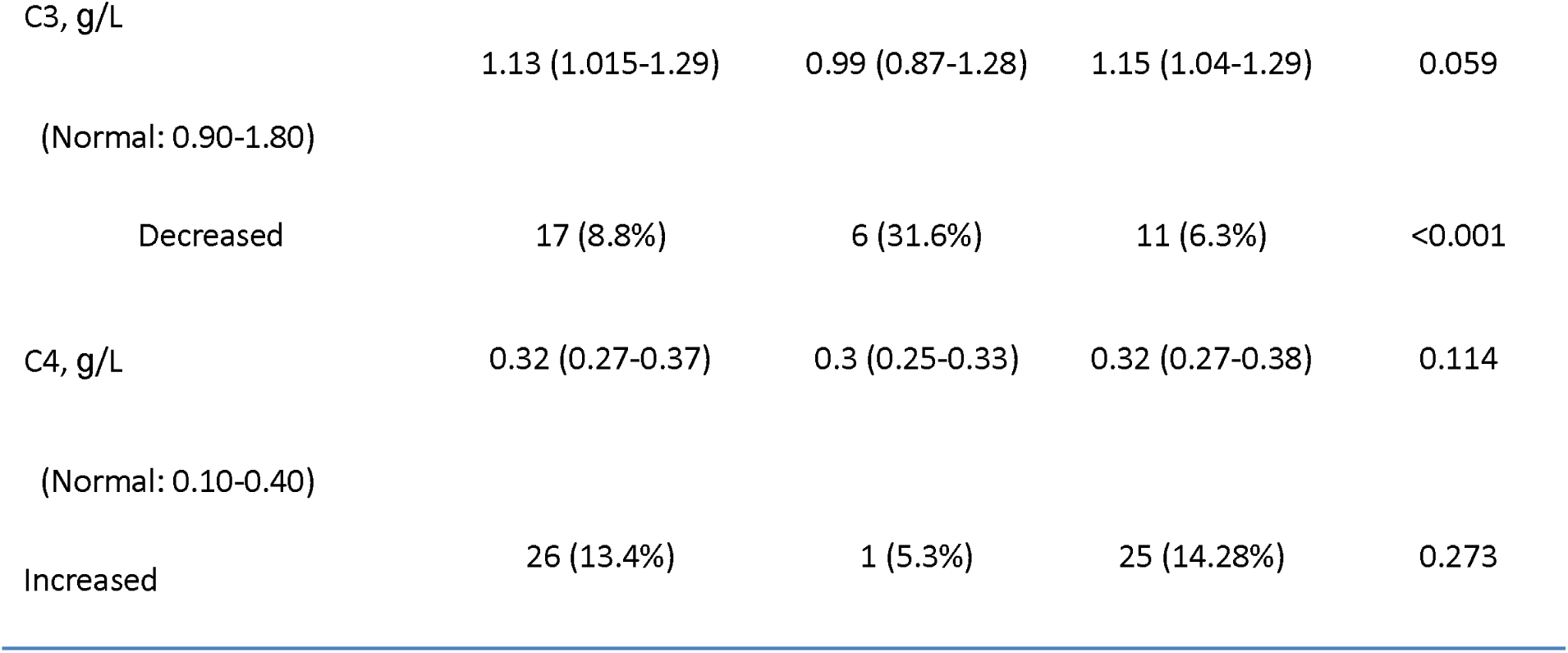
Immunological findings of patients with COVID-19 on admission to hospital

|  |  |  |  |
| --- | --- | --- | --- |
| Cluster of Differentiation (CD) | n=192 | n=19 | n=173 |

|  |  |  |  |  |
| --- | --- | --- | --- | --- |
| CD3, cell/μl | 766.5 (500.5-1055.0) | 411.0<br>(198.0-537.0) | 801.0<br>(561.5-1087.0) | <0.001 |
| (Normal: 690-2540) |  |  |  |  |
| Decreased | 88 (45.8%) | 17 (89.5%) | 71 (41.0%) | <0.001 |
| CD4, cell/μl | 433.5 (283.5-650.75) | 198.0<br>(116.0-340.0) | 468.0<br>(309.5-679.5) | <0.001 |
| (Normal: 410-1590) |  |  |  |  |
| Decreased | 88 (45.8%) | 15 (78.9%) | 73 (42.2%) | 0.002 |
| CD8, cell/μl | 259.5 (162.25-404.0) | 128.0<br>(65.0-182.0) | 217.0<br>(176.0-415.0) | <0.001 |
| (Normal: 190-1140) |  |  |  |  |
| Decreased | 66 (34.4%) | 16 (84.2%) | 51 (29.7%) | <0.001 |
| CD45, cell/μl | 1110.5<br>(749.25-1504.25) | 660.0<br>(435.0-844.0) | 1148.0<br>(797.0-1536.0) | <0.001 |
| (Normal: 900-3500) |  |  |  |  |
| Decreased | 73 (38.0%) | 16 (84.2%) | 57 (32.9%) | <0.001 |
| CD4/8 | 1.695 (1.248-2.293) | 1.78 (1.29-2.15) | 1.68 (1.23-2.32) | 0.828 |
| (Normal: 0.9-3.3) |  |  |  |  |
| Decreased | 12 (6.2%) | 1 (5.3%) | 11 (6.4%) | 0.878 |
| Immunoglobulin | n=194 | n=19 | n=175 |  |
| IGA, g/L | 2.36 (1.8475-3.0925) | 2.18 (1.84-3.68) | 2.38 (1.85-3.04) | 0.775 |
| (Normal: 0.70-4.00) |  |  |  |  |
| Increased | 17 (8.8%) | 4 (21.1%) | 13 (7.4%) | 0.046 |
| IGG, g/L | 11.8 (10.4-13.625) | 12.4 (10.9-13.8) | 11.8 (10.3-13.6) | 0.790 |
| (Normal: 7.00-16.00) |  |  |  |  |
| Increased | 18 (9.3%) | 1 (5.3%) | 17 (9.7%) | 0.525 |
| IGM, g/L | 0.935 (0.69-1.2125) | 0.86 (0.68-0.99) | 0.96 (0.69-11.24) | 0.152 |
| (Normal: 0.40-2.30) |  |  |  |  |
| Increased | 10 (5.2%) | 0 (0.0%) | 10 (5.7%) | 0.602 |
| C3, g/L | 1.13 (1.015-1.29) | 0.99 (0.87-1.28) | 1.15 (1.04-1.29) | 0.059 |
| (Normal: 0.90-1.80) |  |  |  |  |
| Decreased | 17 (8.8%) | 6 (31.6%) | 11 (6.3%) | <0.001 |
| C4, g/L | 0.32 (0.27-0.37) | 0.3 (0.25-0.33) | 0.32 (0.27-0.38) | 0.114 |
| (Normal: 0.10-0.40) |  |  |  |  |
| Increased | 26 (13.4%) | 1 (5.3%) | 25 (14.28%) | 0.273 |

Compared with the non-ICU patients, patients admitted to the ICU had reduced CD3^+^ T cell counts (411.0 vs 801.0) and were more likely to have decreased CD4^+^ T cells (89.5% vs 41.0%), decreased CD8^+^ T cells (84.2% vs 29.7%), decreased CD45^+^leukocyte population (84.2% vs 32.9%) and decreased complement 3 level (31.6% vs 6.3%). By contrast, patients admitted to the ICU were more likely to have increased level of IgA (21.1% vs 7.4%).

### Radiological findings

Of 198 patients who underwent chest computed tomography on admission, 98.5 % cases manifested abnormalities suggestive of pneumonia. The most common patterns on chest computed tomography were ground-glass opacity and bilateral patchy consolidation. Figure 1 illustrates representative findings with different degrees of radiological abnormalities from seven patients.

**Figure 1.**
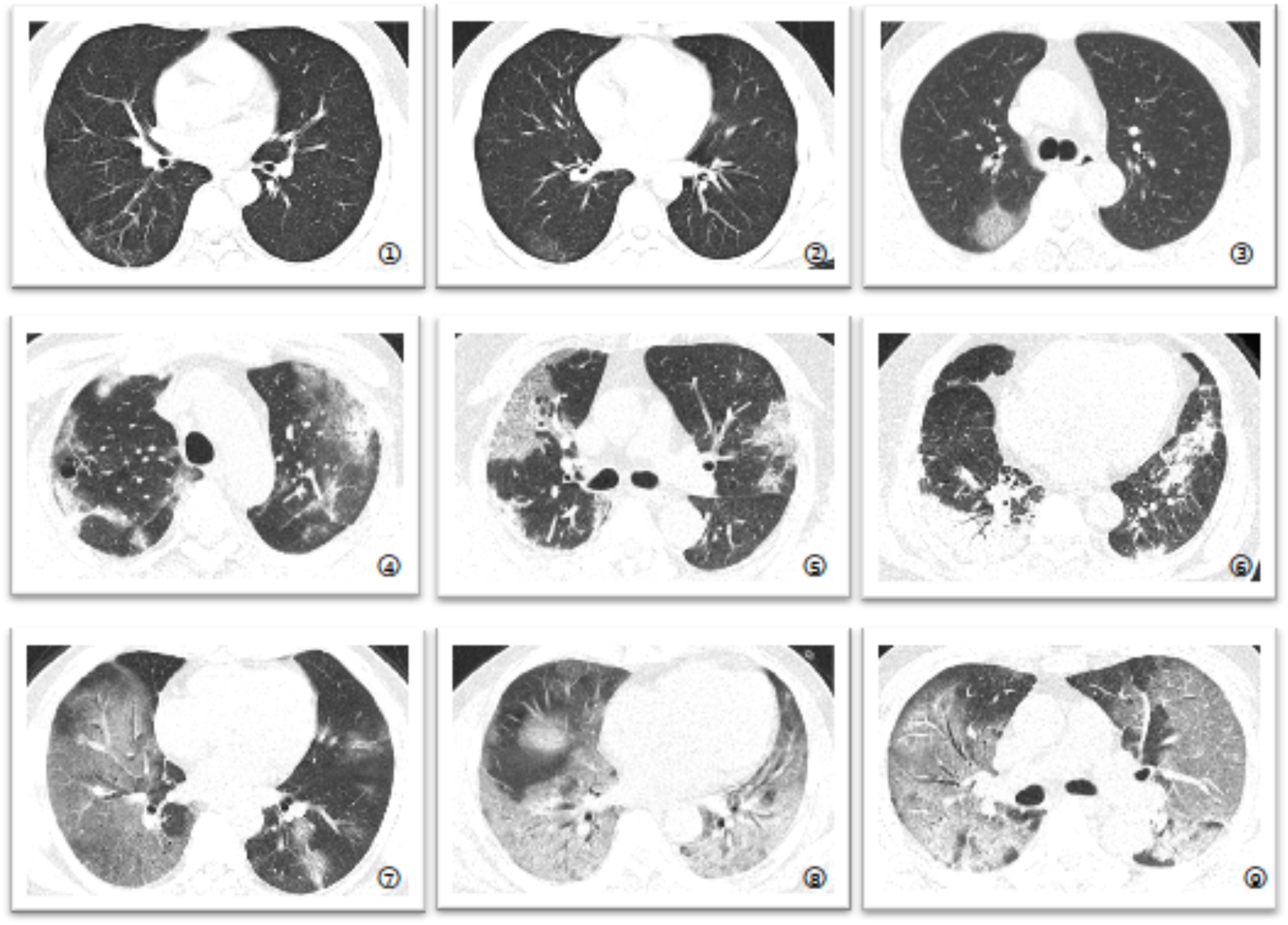
Representative chest CT images from multiple patients. Various radiographic abnormalities are present as follows: 1. Intralobular septal thickening and parenchymal bands in the right lower lobe. 2. A small region of subpleural ground glass opacity with partial consolidation in the right lower lobe. 3. A small region of subpleural consolidation in the posterior right upper lobe. 4. Bilateral multilobular ground glass opacity and partial consolidation, and a pneumatocele in the right upper lobe. 5. Bilateral consolidation in the peripheral regions of the upper lobes, with inter- and intra-lobular septal thickening (crazy paving pattern). 6. Bilateral consolidation in the subpleural region of lower lobes, with parenchymal bands. Air bronchograms are present in the right lower lobe. 7. Diffuse consolidation in the right lower lobe. Ground glass opacity with partial consolidation in multiple lobes of left lung. 8. Bilateral diffuse consolidation with air bronchograms in the lower lobes. Multiple areas of patchy consolidation in the right middle lobe. 9. Bilateral diffuse consolidation with air bronchograms.

### Longitudinal profile of laboratory finding in patients admitted to ICU

To determine the major clinical features that appeared during COVID-19 progression, the dynamic changes in 11 clinical laboratory parameters, including haematological and biochemical parameters, were tracked from day 1 to day 15 after the onset of the disease at 2-day intervals (Fig. 2 &3). ICU patients were subdivided into two subgroups as described in the Methods section: severe and critical. The levels of CRP and procalcitonin in critically ill patients showed a trend for sustained elevation 9 days after admission (Fig. 2A&B). White blood cells and neutrophils counts were higher in critical patients than in severe patients (Fig. 2C&D). Most significantly, critical cases had more severe lymphopenia than severe cases (Fig 2E, < 0.6 × 10^9^/L, P<0.05).

**Figure 2.**
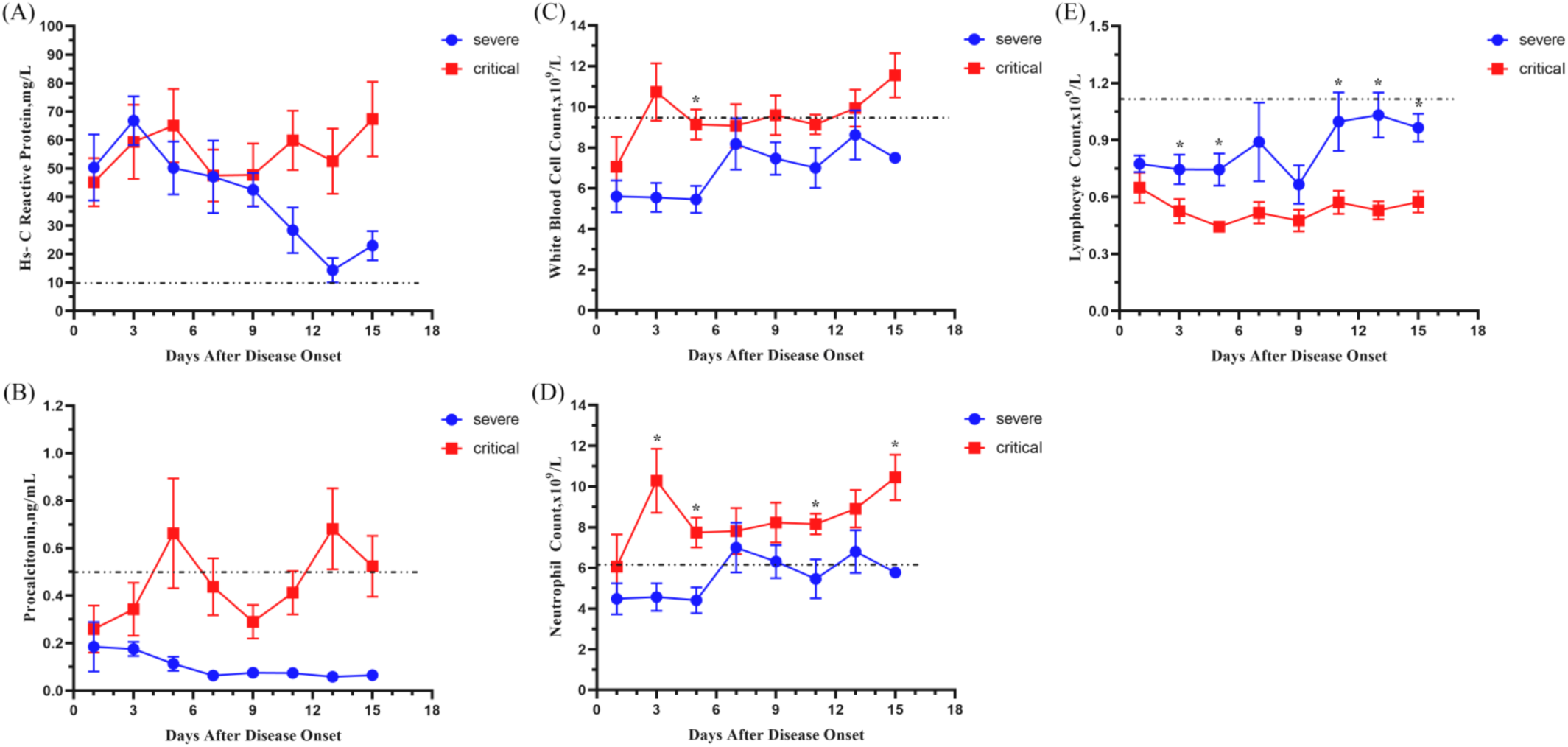
Timeline charts illustrate the laboratory parameters in 19 patients with COVID-19 (9 severe and 10 critical cases) measured every other day based on the days after the onset of illness. Dynamic changes in the following parameters are shown: A) CRP, B) procalcitonin, C) WBC count, D) neutrophil count, E) lymphocyte count. The straight lines in each graph mark the upper (A-D) or lower (E) normal limit of each parameter. *P<0.05, critical group vs severe group at that time point.

Critical cases had significantly more organ dysfunction and coagulation abnormalities than severe cases. Cardiac troponin I and myoglobin was higher in critical patients than in severe cases (Fig. 3A&B), indicating heart injury. The levels of blood urea nitrogen (BUN) and creatinine level were higher in critical cases than in severe cases (Fig. 3C&D), suggesting kidney injury. The level of D-dimer was higher in critically ill than in severely ill (Fig. 3E), however, there was no marked difference in fibrinogen level between these two groups (Fig. 3F).

**Figure 3.**
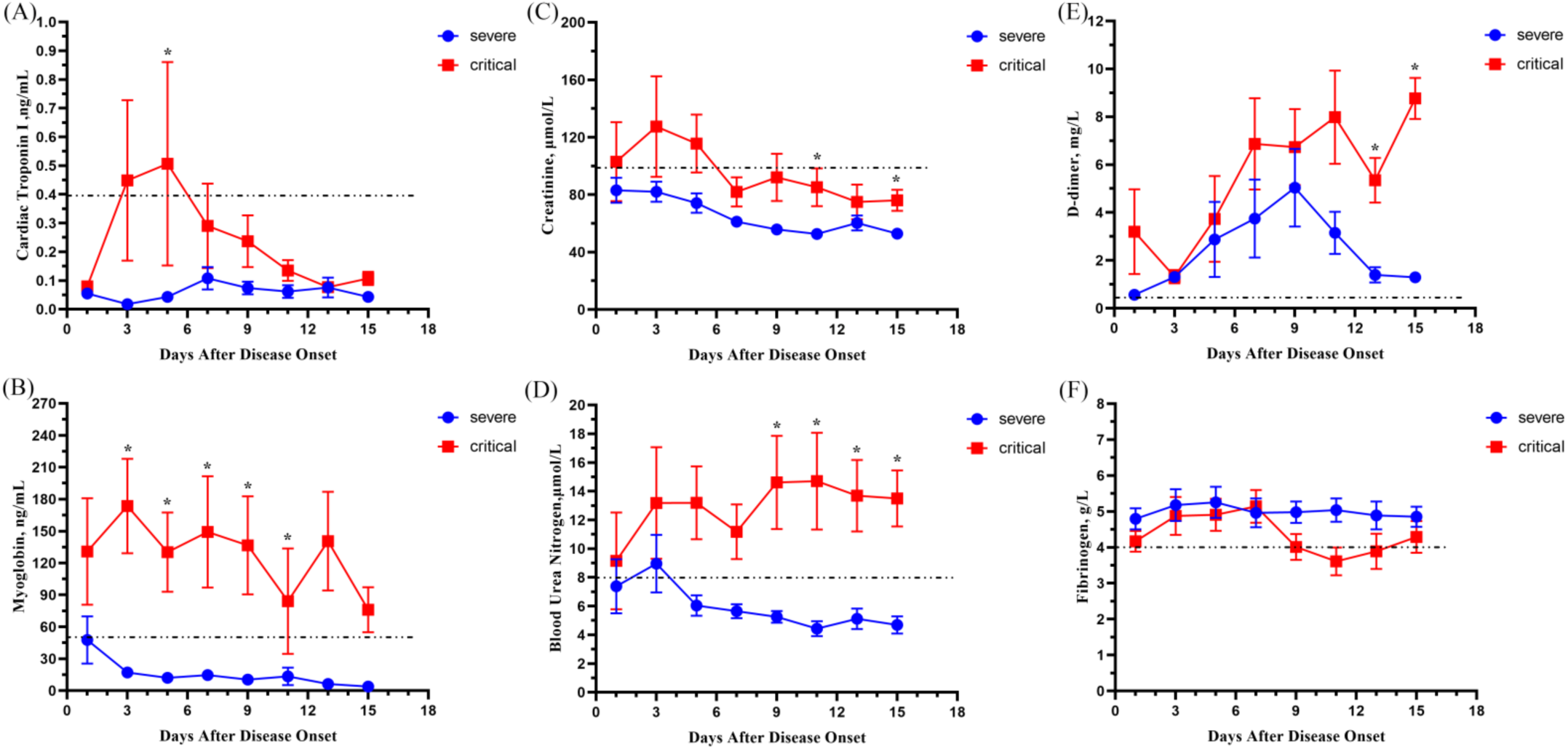
Timeline charts illustrate the laboratory parameters in 19 patients with COVID-19 (9 severe and 10 critical cases) measured every other day based on the days after the onset of illness. Dynamic changes in the following parameters are shown: A) troponin I, B) myoglobin, C) creatinine, D) BUN, E) D-dimer, F) fibrinogen. The straight lines in each graph mark the upper normal limit of each parameter. *P<0.05, critical group vs severe group at that time point.

## Discussion

In this single centre cohort of confirmed COVID-19 cases in Shanghai, the median duration of symptoms from disease onset to hospital admission was 4 days. Compared to the previously reported duration of 7-10 days between symptom onset and hospitalization in Wuhan ^3 5 6^, patients in Shanghai received earlier in-ward observation and medical service, most likely due to improved monitoring and increased awareness of the outbreak. As a result, our data likely were recorded closer to the onset date of the disease, which may explain some of the differences between the results reported from Wuhan and Shanghai.

In terms of patient’s characteristics, we observed a comparatively equal male to female ratio in this study, which differs from the data reported in Wuhan ^3 5 6^. This may be because most of these initial cases in Shanghai had a history of exposure in Wuhan (63.6%), and the disease thereafter appeared in a family cluster pattern. Almost half of the patients in Shanghai were under 50 years old, in contrast with the predominantly older patient population reported in Wuhan. In addition, 69 (34.8%) patients in our cohort had pre-existing comorbidities. Whether the disease is more likely to manifest in patients with comorbidities and/or older patients remains unclear and requires further study.

Regarding laboratory evaluation, low or normal levels of white blood cell (WBC) and neutrophil counts were present in COVID-19 patients upon admission. ESR and CRP levels were elevated in most patients. Although procalcitonin level was increased in over one-third (28.4%) of the patients, the median 0.03 (0.02-0.06) was close to normal range. These results are similar with that of two Wuhan cohort studies already published ^3 5^. We further observed that serum lactate levels were above the normal upper range in the majority of patients (83.7%), indicating the likely presence of metabolic acidosis. The sensitivity of LACT in COVID-19 infection is similar to ESR and CRP.

In addition, we observed a marked reduction in T and TH subtype cells in infected patients, suggesting that SARS-Cov-2 infection may impair cellular immunity. It is known that MERS-CoV is able to infect both CD4^+^ and CD8^+^ primary human T cells and, upon infection induces T cell apoptosis in vitro ^7^. While the cell surface receptor for SARS-Cov-2 has now been identified as ACE2 ^8^, other receptors may also exist on immune cells to bind to coronavirus ^9-11^. As there is no proven antiviral treatment yet available, strategies to enhance the immune system may be considered. Since the most common haematological changes in COVID-19 patients were lymphopenia and immunodeficiency, we postulate that hematopoietic growth factors such as G-CSF, by mobilizing endogenous blood stem cells and endogenous cytokines, may represent a potential haematological treatment for COVID-19 patients ^12^.

Interestingly, a noticeable increase of glutathione reductase (GR) level occurred in 40.2% of the COVID-19 patients in our cohort, while ALT and AST are normal in most patients. GR is an essential enzyme that recycles oxidized glutathione back to the reduced form ^13^. GR is known to participate in an oxidative defence system required for effective immune responses against bacteria ^14 15^. Whether GR is involved in host defence systems against viruses such as SARS-Cov-2 remains to be determined.

We observed that 65.4% of COVID-19 patients had decreased serum calcium levels. Calcium influx regulates both clathrin-mediated and clathrin-independent endocytosis during viral infection, thus constituting a key mechanism for regulation of influenza A virus internalization and infection ^16^. Based on this theory, calcium channel blockers, including amlodipine, verapamil, and diltiazem, as well as BAPTA-AM, have been proven effective in inhibiting IAV replication in a dose-dependent manner in a canine kidney cell model ^17^. Moreover, diltiazem may have both prophylactic and therapeutic effects in IAV treatment according to both ex vivo and in vivo testing ^17^. We therefore propose the hypothesis that Ca^2+^ levels and/or Ca^2+^ channels may play a role in endocytosis and infection of SARS-Cov-2. Further studies are needed to characterize the functional importance of this potential pathway.

The patients admitted to the ICU were more likely to be older, male, with temperature over 38.5°C, symptom of dyspnoea, underlying cardiovascular disease, and longer waiting period from onset of symptom to hospital admission, compared to those not admitted to the ICU. This suggests that age, sex, high fever, admission time and co-morbidity are risk factors for disease severity. Compared with non-ICU patients, patients who received ICU care had numerous laboratory abnormalities. These abnormalities suggest that SARS-Cov-2 infection can be associated with cellular immune deficiency, coagulation activation, myocardial injury, hepatic injury and kidney injury. These laboratory abnormalities are similar to those previously observed in patients with MERS-Cov and SARS-Cov infection ^18 19^.

The longitudinal profile of laboratory findings was dynamically tracked for those patients requiring ICU care. When these patients were subdivided into severe and critical ill subgroups, we observed that white blood cell count, neutrophil count, D-dimer, BUN, creatinine, myoglobulin and troponin I levels were higher in critical cases than in severe cases. Neutrophilia may be associated with secondary infection, while coagulation activation could be related to sustained inflammatory responses. Acute heart and kidney injury could be related to direct effects of the virus or possibly hypoxia ^9 20^.

Our study has several limitations. First, although we obtained data from the initial 198 patients with laboratory-confirmed COVID-19 in Shanghai, the cohort is still relatively small. More patients need to be analysed to provide a comprehensive and precise description of the spectrum of disease associated with this infection. Secondly, our study reports primarily baseline results from patients upon hospital admission, and more longitudinal data regarding disease progression and clinical outcomes will require further collection and study. Despite these limitations, our study provides data from the first large cohort outside Wuhan and adds important laboratory information to the rapidly accumulating body of information about COVID-19. These results will assist in multi-centre monitoring of the disease.

In conclusion, COVID-19 affects a wide-range of patients, from youth to the elderly. Fever is consistently the most common symptom of onset, but multiple other clinical manifestations occur, including a spectrum form T cell deficiency to symptoms of digestive system involvement. Older age, male, fever over 38.5°C, symptom of dyspnoea, the presence of underlying cardiovascular disease, and longer waiting period from onset of symptom to hospital admission are risk factors associated with the severity of disease.

## Data Availability

The data that support the findings of this study are available from the corresponding authors upon request. Participant data are without names and identifiers, and they will be made available after approval from the corresponding author and National Health Commission. After publication of study findings, the data will be available for others to request. The research team will provide an email address for communication once the data are approved for sharing.

## Acknowledgments

We thank Dr. Suling Li (Lewis University) for providing guidance on the analysis of data, and Dr. Duan Zhou (Longhua Hospital, Shanghai University of Traditional Chinese Medicine) for assistance in the preparation of this manuscript.

## Ethical approval

The study was approved by the ethics committee of Shanghai Public Health Clinical Centre, Shanghai, China

## Footnotes

Contributors: MC, DZ, Y-FL, YL, HX, ZY, JS and MZ enrolled study subjects and collected data. YW, XZ and PL, participated in the study design and conception. YL, YW, CZ, and YD performed statistical analysis of the data. YW, XZ, SD wrote the manuscript. All authors reviewed and approved the final manuscript as submitted and agree to be accountable for all aspects of the work.

## Funding information

This research was funded by the First-rate University and Discipline Construction Project of Fudan University (IDF162005) and the Key Scientific Research Projects on 2019-nCoV of Shanghai Public Health Clinical Centre (2020YJKY01), the Special Emergency Project for the Prevention and Treatment of COVID-19 with Traditional Chinese Medicine in Shanghai(2020NCP001), and the 2018-2020 Three-year Action Plan for Traditional Chinese Medicine Further Development in Shanghai (ZY (2018-2020) CCCX-2002-04).

## Disclaimer

The funders did not play any role in the design of the study, data collection, analysis and interpretation of data, and in writing the manuscript.

Competing interests: ZX holds research grants from Shanghai municipal government. HL holds research grant from Fudan University and from Shanghai public health clinical centre. All the other authors have no disclosures.

## Patient consent for publication

Patient consent was waived.

## Notes

### Competing Interest Statement

The authors have declared no competing interest.

### Clinical Trial

Histologic features

